# Physical activity and risk of breast and endometrial cancers: a Mendelian randomization study

**DOI:** 10.1101/19005892

**Authors:** Hansjörg Baurecht, Michael Leitzmann, Tracy O’Mara, Deborah J Thompson, the collaborators of the Endometrial Cancer Association Consortium, Alexander Teumer, Sebastian E. Baumeister

## Abstract

**Importance:** The causality of the association between physical activity and risk of breast and endometrial cancers is uncertain because available evidence is based exclusively on observational studies, which are potentially susceptible to confounding and reverse causation.

**Objective:** To investigate whether increased physical activity is causally associated with decreased risk of breast and endometrial cancers, using a two-sample Mendelian randomization study design.

**Design, Setting, and Participants:** Genome-wide association studies of physical activity, breast cancer, and endometrial cancer, published up to April 31, 2019, were identified using PubMed and the GWAS catalog. Twelve single nucleotide polymorphisms (SNP) known at P < 5 × 10^−8^ to be associated with accelerometer-assessed or self-reported physical activity served as instrumental variables. Genetic summary data from four large consortia provided SNP-outcome associations [Breast Cancer Association Consortium; Discovery, Biology and Risk of Inherited Variants in Breast Cancer Consortium; Endometrial Cancer Association Consortium].

**Main Outcomes and Measures:** The primary outcomes were risk of breast cancer and risk of endometrial cancer. Secondary outcomes were estrogen receptor positive (ER+) and ER-breast cancers. Odds ratios (ORs) and 95% confidence intervals (CIs) per mean acceleration in milli-gravities of accelerometer-assessed physical activity and per one standard deviation (1-SD) increase in metabolic-equivalent (MET)-minutes/week of self-reported moderate-to-vigorous physical activity were computed using the inverse variance weighted method. A series of sensitivity analyses addressed the potential impact of heterogeneity, pleiotropy, and outliers.

**Results:** Summary data were available for 122,977 breast cancers and 12,270 endometrial cancers. Genetic predisposition to increased accelerometer-assessed physical activity was associated with lower risk of breast and endometrial cancers. The associations (ORs [95% CI] per 1-SD increase in mean acceleration) were 0.88 (0.85-0.91) for breast cancer and 0.90 (0.83-0.97) for endometrial cancer. In addition, genetic predisposition to increased accelerometer-assessed physical activity was associated with lower risk of ER+ breast cancer. We found no evidence for an association between genetic predisposition to self-reported physical activity and risk of total breast cancer, breast cancer subtypes, or endometrial cancer.

**Conclusion and Relevance:** This first Mendelian randomization study shows that objectively-assessed physical activity plays a causal role in protecting against breast and endometrial cancers.

## Introduction

Breast cancer is the most common female malignancy and endometrial cancer is the sixth most common female malignancy in high-income countries ^1^. Given the large burden of breast and endometrial cancers, it is imperative to identify modifiable risk factors for effective prevention. For example, several non-genetic risk factors have been established for endometrial cancer, including obesity, insulin resistance, and certain reproductive factors ^2^. Although the causality of some of those factors, such as obesity ^2,3^, is well accepted, relations with other potential risk factors remain uncertain.

The association between physical activity and risk of breast and endometrial cancers has received considerable attention in recent years, with umbrella reviews and meta-analyses of observational studies providing evidence for protective associations with both breast cancer ^4-6^ and endometrial cancer ^2,6,7^. Those associations have been confirmed by reports of the World Cancer Research Fund ^3^ and the 2018 U.S. Physical Activity Guidelines Advisory Committee ^8^. All of the evidence underpinning physical activity guidelines has come from observational studies linking physical activity data to cancer risk ^8^. However, observational studies often cannot distinguish true, causal associations from artefactual associations that may be a consequence of confounding or reverse causation ^9^.

By comparison, randomized controlled trials can provide unequivocal evidence for causality, but are often infeasible given the long latency periods for many cancers ^10^. Genetic variants can be useful to infer causality because they are present from birth and are therefore unlikely to be confounded by environmental factors ^11^. Mendelian randomization is a method that uses genetic variants as instrumental variables to uncover causal relationships in the presence of observational study bias ^11^. Previous Mendelian randomization studies have confirmed obesity as a causal risk factor in the development of both breast and endometrial cancers ^12-14^. In the current study, we performed two-sample Mendelian randomization analyses to assess the causal association of physical activity with risk of breast and endometrial cancers.

## Methods

The current study design comprised of three components: (1) identification of genetic variants to serve as instrumental variables for physical activity; (2) acquisition of summary data for the genetic instruments from genome-wide association studies on physical activity; (3) acquisition of instrumenting SNP-outcome summary data from genome-wide association studies of breast cancer and endometrial cancer.

### Selection of Instrumental Variables for Physical Activity

We searched the GWAS catalog ^15^ and PubMed for studies published up to April 31, 2019, to identify single-nucleotide polymorphisms (SNPs) associated with physical activity (eTable 1). We used a previous genome-wide association study of 91,084 UK Biobank participants that identified three SNPs associated with accelerometer-assessed physical activity (mean acceleration in milli-gravities) at genome-wide significance (P < 5×10^−8^) ^16^. We also used nine SNPs previously associated with 1-standard deviation (SD) increase in metabolic-equivalent (MET)-minutes/week of self-reported moderate-to-vigorous physical activity at genome-wide significance in 377,234 individuals ^16^. The estimated ‘Chip-heritability’ for accelerometer-assessed and self-reported physical activity was 14% and 5%, respectively ^16^.

### Outcome Data

We used publicly available summary statistics data for genetic associations with breast cancer and endometrial cancer ^17,18^. Specifically, the meta-analysis of genome-wide association studies of the Breast Cancer Association Consortium and the Discovery, Biology and Risk of Inherited Variants in Breast Cancer Consortium included data from 122,977 female cases and 105,974 female controls of European descent ^17^. Cases were subtyped according to estrogen receptor (ER) positive (n = 69,501) and ER negative status (n= 21,468). The Endometrial Cancer Association Consortium provided summary statistics for from their recent genome-wide association meta-analysis ^18^ with the UK Biobank strata removed (total dataset 12,270 endometrial cancer cases and 46,126 controls of European descent) (eTable 2). The study consortia data for breast cancer did not include the UK Biobank.

## Statistical analysis

The principal analysis was conducted using the inverse-variance weighted (IVW) method, under a fixed effects model, alongside other methods to overcome violations of specific instrumental variable assumptions: weighted median, MR-Egger and MR-Pleiotropy RESidual Sum and Outlier (MR-PRESSO) ^19,20^. Results are presented as odds ratios (OR) and 95% confidence intervals (CIs) per mean acceleration in milli-gravities of accelerometer-assessed physical activity and per 1-SD increase in MET-minutes/week of self-reported moderate-to-vigorous physical activity. We used the Cochran Q statistic to test for heterogeneity ^19,21^. Higher levels of heterogeneity in the estimated effects from each SNP indicate potential pleiotropy, which potentially introduces bias in Mendelian randomization estimation ^21^. We performed leave-one-out analyses for accelerometer-based and self-reported activity to address potential heterogeneity. We assessed potential directional pleiotropy by testing the intercepts of MR-Egger models ^19^. Analyses were performed using the TwoSampleMR (version 0.4.22) ^20^ and MR-PRESSO (version 1.0) packages in R (version 3.5.3).

## Results

Mendelian randomization analyses revealed inverse associations of accelerometer-assessed physical activity with both breast and endometrial cancers. Specifically, genetically predicted accelerometer-assessed physical activity was associated with a 12% lower odds of breast cancer (IVW OR per mean acceleration in milli-gravities: 0.88; 95% CI: 0.85-0.91, P=5.7×10^−16^) (Table 1). Analyses according to breast cancer subtype showed an inverse relationship of accelerometer-assessed physical activity to ER+ breast cancer (IVW OR: 0.86; 95% CI: 0.83-0.89; P=1.7×10^−15^) but no association with ER-breast cancer (IVW OR 0.99; 95% CI: 0.94-1.05; P=.788). Genetically predicted accelerometer-assessed physical activity was also associated with a 10% lower odds of endometrial cancer (IVW OR: 0.90; 95% CI: 0.83-0.97, P=6.5×10^−3^) (Table 1). By comparison, genetically predicted self-reported physical activity showed no associations with total breast cancer, ER+ breast cancer, ER-breast cancer, or endometrial cancer (Table 2).

**Table 1.**
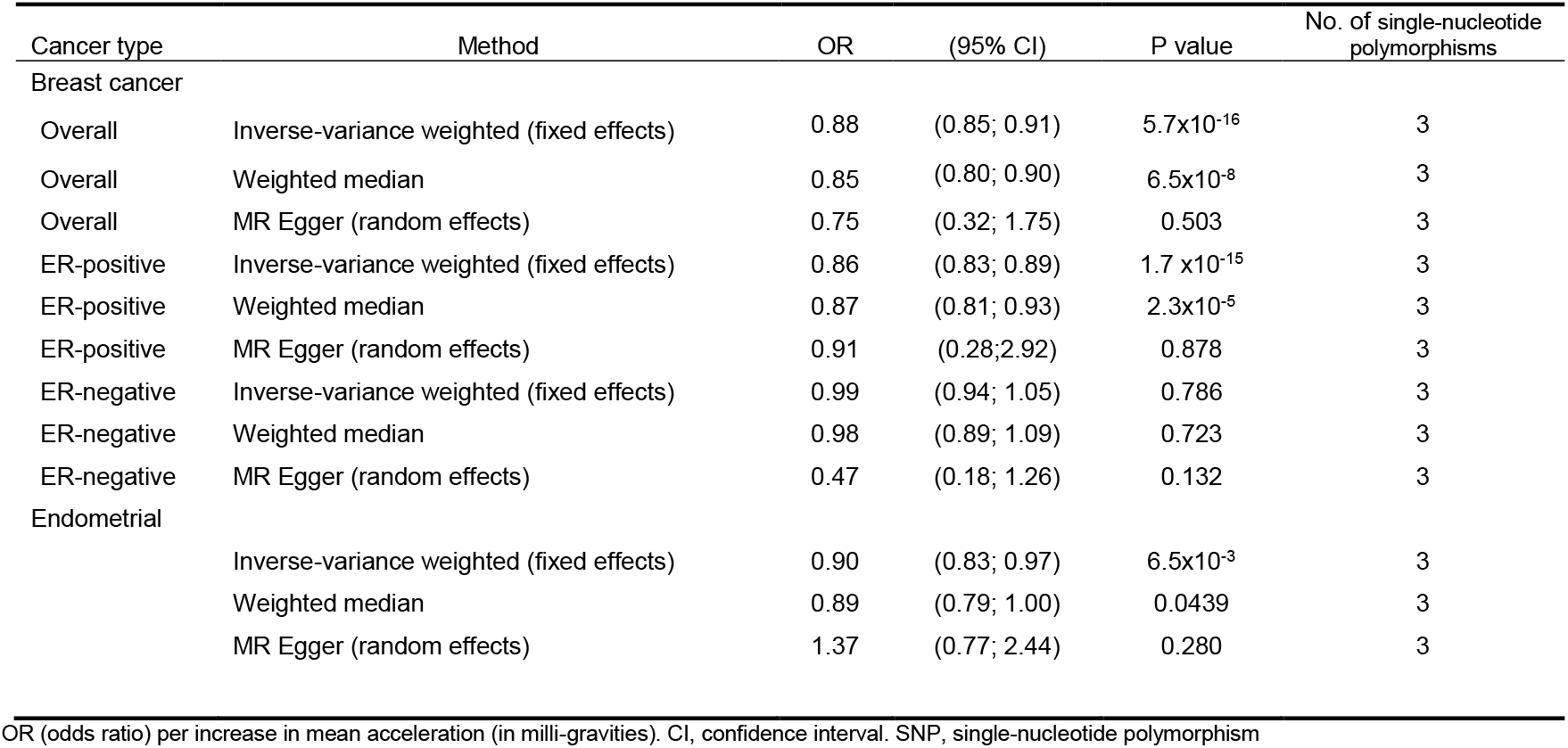
Mendelian randomization estimates between accelerometer-assessed physical activity and cancer risk

**Table 2.** Mendelian randomization estimates between self-reported physical activity and cancer risk

| Cancer type | Method | OR | (95% CI) | P value | No. of single-nucleotide polymorphisms |
| --- | --- | --- | --- | --- | --- |
| Breast cancer |  |  |  |  |  |
| Overall | Inverse-variance weighted (fixed effects) | 0.94 | (0.71; 1.27) | 0.703 | 9 |
| Overall | Weighted median | 1.10 | (0.69; 1.74) | 0.690 | 9 |
| Overall | MR Egger (random effects) | 2.02 | (0.43; 9.54) | 0.374 | 9 |
| Overall | MR PRESSO | 0.94 | (0.62; 1.44) | 0.797 | 9 |
| ER-positive | Inverse-variance weighted (fixed effects) | 0.97 | (0.69; 1.38) | 0.872 | 9 |
| ER-positive | Weighted median | 1.18 | (0.71; 1.97) | 0.517 | 9 |
| ER-positive | MR Egger (random effects) | 1.55 | (0.27; 8.90) | 0.6215 | 9 |
| ER-positive | MR PRESSO | 0.97 | (0.62; 1.52) | 0.903 | 9 |
| ER-negative | Inverse-variance weighted (fixed effects) | 0.85 | (0.50; 1.46) | 0.557 | 9 |
| ER-negative | Weighted median | 0.71 | (0.32; 1.57) | 0.394 | 9 |
| ER-negative | MR Egger (random effects) | 3.79 | (0.41; 35.04) | 0.241 | 9 |
| ER-negative | MR PRESSO | 0.85 | (0.46-1.58) | 0.624 | 9 |
| Endometrial |  |  |  |  | 9 |
|  | Inverse-variance weighted (fixed effects) | 0.90 | (0.43; 1.87) | 0.780 | 9 |
|  | Weighted median | 0.51 | (0.14; 1.82) | 0.299 | 9 |
|  | MR Egger (random effects) | 30.60 | (0.15; 6416.11) | 0.210 | 9 |
|  | MR PRESSO | 0.56 | (0.17; 1.77) | 0.357 | 7 |

MR PRESSO, MR Pleiotropy RESidual Sum and Outlier. OR (odds ratio) per one standard increase in metabolic-equivalent (MET)-minutes/week. CI, confidence interval. SNP, single-nucleotide polymorphism

We performed sensitivity analyses to examine potential pleiotropy and outliers. The Cochran Q statistics indicated horizontal pleiotropy for the associations of accelerometer-assessed physical activity with total breast cancer and ER+ breast cancer, but not with ER-breast cancer and endometrial cancer (eTables 3). Analyses also suggested horizontal pleiotropy for the relations of self-reported activity to overall breast cancer and endometrial cancer (eTables 3). Results from the leave-one-out analyses of accelerometer-assessed and self-reported activity did not substantially alter the results (eTables 4 and 5). There was no indication of directional pleiotropy (all P >.21, eTable 6). MR-PRESSO did not detect any outlier for models of self-reported physical activity in relation to breast cancer, but it detected two outliers (rs2854277, rs429358) for models of self-reported physical activity in relation to endometrial cancer (Table 1). Due to the limited number of available SNPs, MR-PRESSO was not performed for accelerometer-assessed physical activity.

## Discussion

To the best of our knowledge, the current study represents the first Mendelian randomization analysis of genetically determined and objectively assessed physical activity in relation to cancer risk. Our investigation provides strong support for causal inverse associations between accelerometer-assessed physical activity and risks of breast and endometrial cancers. Specifically, our analyses show that an increase in genetically determined, accelerometer-assessed physical activity is associated with a 12% lower risk of breast cancer and a 10% lower risk of endometrial cancer. We also detected a 14% risk reduction for ER+ breast cancer associated with an increase in genetically determined, accelerometer-assessed physical activity. By comparison, we found no association between genetically determined, accelerometer-assessed physical activity and ER-breast cancer. In addition, we found no associations between genetically determined, self-reported physical activity and risks of breast or endometrial cancers.

Our findings are largely consistent with previous prospective observational investigations of physical activity and risk of breast and endometrial cancers ^4-8^. A large pooled analysis of 10 European and US cohort studies including 35,178 breast cancer cases reported a risk reduction of 10% comparing high versus low levels of self-reported physical activity (hazard ratio: 0.90; 95%: 0.87-0.93) ^22^. Similarly, the most comprehensive available meta-analysis of self-reported physical activity and breast cancer comprising 38 prospective studies and 116,304 breast cancer cases found that the highest compared to the lowest level of physical activity was associated with a 12% risk reduction (relative risk: 0.88; 95%: 0.85-0.90) ^5^. In addition, high versus low self-reported physical activity was associated with a 11% reduced risk of ER+ breast cancer (relative risk: 0.89; 95% CI: 0.83-0.95) ^5^. A pooled analysis of nine cohort studies including 5,347 endometrial cancer cases reported a risk reduction of 21% (hazard ratio: 0.79; 95 CI: 0.68-0.92) for high versus low self-reported physical activity ^22^. A recent meta-analysis found a 20% reduction in risk of endometrial cancer when comparing the highest versus the lowest amounts of self-reported physical activity (relative risk: 0.80; 95 CI: 0.75-0.85) ^7^. Of note, although our Mendelian randomization estimates are similar in magnitude to those obtained from pooled analyses and meta-analyses of previous observational studies, Mendelian randomization investigations generally provide estimates of cumulative, long term exposure, whereas risk estimates from observational studies more typically reflect recent past or current levels of physical activity ^23^.

Numerous investigations have evaluated physical activity in relation to breast and endometrial cancers but previous studies have relied exclusively on observational study designs. Therefore, it has been difficult to discern whether associations between physical activity and cancer risk are truly causal ^9^. The ongoing debate about whether physical activity causally protects from cancer has in part been inconclusive due to a lack of instrumental variables ^9^. The current Mendelian randomization study closes this gap by utilizing newly available instrumental variables, thereby improving causal inference and overcoming potential biases of traditional observational studies ^11^.

Mendelian randomization studies have shown that physical activity lowers the risk of adiposity ^24^ and that obesity increases risks of breast and endometrial cancers ^12-14,25^, which led us to hypothesize that physical activity causally acts as a protective factor for breast and endometrial cancers. The validity of the causal estimates derived from Mendelian randomization approaches requires several assumptions to be satisfied ^11^. When the genetic variants selected as instrumental variables are unassociated with potential confounders and are not linked to the outcome via any alternative pathway, Mendelian randomization analysis provides evidence for rejecting the sharp causal null hypothesis of no effect of the exposure of interest ^11,26^. We selected the most significant independent SNPs as instrumental variables such that all SNPs were robustly associated with physical activity. However, although we used the largest genome-wide association study available, only few genome-wide significant instrumenting SNPs were available, leading to wide CIs of the risk estimates.

A major assumption underlying the validity of our Mendelian randomization analysis was that the genetic variant affects breast or endometrial cancer only through physical activity. We conducted a large range of statistical analyses to rule out potential pleiotropy and we applied methods that are particularly robust to horizontal pleiotropy. Although horizontal pleiotropy biases Mendelian randomization estimates, vertical pleiotropy (where the instrument affects a mediator along the causal pathway) does not ^11,19^. For example, although it is often assumed that obesity mediates the inverse association between physical activity and cancer risk (vertical pleiotropy), obesity could also confound that association by rendering physical activity difficult to perform (horizontal pleiotropy) ^9^. We performed sensitivity analyses to address horizontal pleiotropy and found that the exclusion of potentially pleiotropic SNPs did not alter our main findings.

In order for the two-sample Mendelian randomization method to be valid, the two analytic samples should derive from the same underlying population. The discovery genome-wide association study of physical activity consisted of UK Biobank participants of European descent, aged 40 to 70 years ^16^. Data on sex-specific SNP-physical activity associations were unavailable. By comparison, SNP-outcome associations were derived from female samples with an age range broader than 40 to 70 years. Given the limited age range of the UK Biobank and inclusion of European ancestry individuals only, our results may not be generalizable to other age groups or ancestral populations. Therefore, replication of our findings in other age groups and non-European populations is warranted. By using non-specific effects, our analyses assumed that the effects of SNPs on physical activity are not sex-specific and do not vary by age. We expect sex differences were small for the majority of associations. Thus, our Mendelian randomization analysis estimated the risks of breast and endometrial cancers resulting from “life-long” exposure to physical activity-increasing SNPs, which is the effect of the genetic variant determined at conception and assumes that the association between the genetic variants accumulates over time and does not change with age ^23^. However, this may not be an entirely tenable assumption given that the heritability of physical activity has been shown to decrease with age ^27^.

Because available data on SNPs associated with physical activity were limited to moderate-to-vigorous physical activity, we were unable to consider specific types, durations, or frequencies of physical activity. Our analyses showed that accelerometer-assessed physical activity was related to reduced risks of breast and endometrial cancers, whereas self-reported physical activity yielded null associations. One explanation for this finding is that objective measures of physical activity show less random variation than do subjective measures ^28^, which may have generated more precise estimates of accelerometer-assessed as opposed to self-reported physical activity. Furthermore, because some genetic loci for self-reported physical activity are also related to cognitive function, self-reported physical activity measures may be prone to information bias ^16,29^. In contrast, associations based on accelerometer-assessed physical activity are unrelated to cognitive performance, which essentially rules out any impact cognitive biases could have had on our results ^16,29^.

In conclusion, our Mendelian randomization study suggests that increased physical activity is causally related to decreased risk of breast and endometrial cancers. Our results, in combination with previous literature, provide strong evidence that increased physical activity plays an important role in the prevention of breast cancer and endometrial cancer. Future research may identify if the effects of physical activity vary with age to identify critical time windows for designing prevention strategies.

## Data Availability

Genome-wide association study summary data for physical activity and breast cancer can be found at https://drive.google.com/drive/folders/1p2-aKT6GgOv4425yaIcvO0nN30O4mpPh; https://doi.org/10.5287/bodleian:yJp6zZmdj. Summary data of the Endometrial Cancer Association Consortium was provided by Dr O’Mara.

## Acknowledgments

The authors thank the authors of the physical activity genome-wide association study; the Breast Cancer Association Consortium; the Discovery, Biology and Risk of Inherited Variants in Breast Cancer Consortium, and the Endometrial Cancer Association Consortium for providing the summary data. HB and SB had full access to all the data in the study and take responsibility for the integrity of the data and the accuracy of the data analysis.

## References

1. Bray F, Ferlay J, Soerjomataram I, Siegel RL, Torre LA, Jemal A. Global cancer statistics 2018: GLOBOCAN estimates of incidence and mortality worldwide for 36 cancers in 185 countries. CA Cancer J Clin. 2018;68(6):394–424.

2. Raglan O, Kalliala I, Markozannes G, et al. Risk factors for endometrial cancer: An umbrella review of the literature. Int J Cancer. 2018.

3. World Cancer Research Fund International, American Insitute for Cancer Research. Diet, nutrition, physical activity and cancer: a global perspective. third expert report. 2018.

4. Neilson HK, Farris MS, Stone CR, Vaska MM, Brenner DR, Friedenreich CM. Moderate-vigorous recreational physical activity and breast cancer risk, stratified by menopause status: a systematic review and meta-analysis. Menopause. 2017;24(3):322–344.

5. Pizot C, Boniol M, Mullie P, et al. Physical activity, hormone replacement therapy and breast cancer risk: A meta-analysis of prospective studies. Eur J Cancer. 2016;52:138–154.

6. Rezende LFM, Sa TH, Markozannes G, et al. Physical activity and cancer: an umbrella review of the literature including 22 major anatomical sites and 770 000 cancer cases. Br J Sports Med. 2018;52(13):826–833.

7. Schmid D, Behrens G, Keimling M, Jochem C, Ricci C, Leitzmann M. A systematic review and meta-analysis of physical activity and endometrial cancer risk. Eur J Epidemiol. 2015;30(5):397–412.

8. McTiernan A, Friedenreich CM, Katzmarzyk PT, et al. Physical Activity in Cancer Prevention and Survival: A Systematic Review. Med Sci Sports Exerc. 2019;51(6):1252–1261.

9. Wade KH, Richmond RC, Davey Smith G. Physical activity and longevity: how to move closer to causal inference. Br J Sports Med. 2018;52(14):890–891.

10. Yarmolinsky J, Wade KH, Richmond RC, et al. Causal Inference in Cancer Epidemiology: What Is the Role of Mendelian Randomization? Cancer epidemiology, biomarkers & prevention : a publication of the American Association for Cancer Research, cosponsored by the American Society of Preventive Oncology. 2018;27(9):995–1010.

11. Burgess S, Foley CN, Zuber V. Inferring Causal Relationships Between Risk Factors and Outcomes from Genome-Wide Association Study Data. Annu Rev Genomics Hum Genet. 2018;19:303–327.

12. Pierce BL, Kraft P, Zhang C. Mendelian randomization studies of cancer risk: a literature review. Curr Epidemiol Rep. 2018;5(2):184–196.

13. Shu X, Wu L, Khankari NK, et al. Associations of obesity and circulating insulin and glucose with breast cancer risk: a Mendelian randomization analysis. Int J Epidemiol. 2018.

14. O’Mara TA, Glubb DM, Kho PF, Thompson DJ, Spurdle AB. Genome-Wide Association Studies of Endometrial Cancer: Latest Developments and Future Directions. Cancer epidemiology, biomarkers & prevention : a publication of the American Association for Cancer Research, cosponsored by the American Society of Preventive Oncology. 2019.

15. Buniello A, MacArthur JAL, Cerezo M, et al. The NHGRI-EBI GWAS Catalog of published genome-wide association studies, targeted arrays and summary statistics 2019. Nucleic Acids Res. 2019;47(D1):D1005–d1012.

16. Klimentidis YC, Raichlen DA, Bea J, et al. Genome-wide association study of habitual physical activity in over 377,000 UK Biobank participants identifies multiple variants including CADM2 and APOE. Int J Obes (Lond). 2018;42(6):1161–1176.

17. Michailidou K, Lindstrom S, Dennis J, et al. Association analysis identifies 65 new breast cancer risk loci. Nature. 2017;551(7678):92–94.

18. O’Mara TA, Glubb DM, Amant F, et al. Identification of nine new susceptibility loci for endometrial cancer. Nat Commun. 2018;9(1):3166.

19. Hemani G, Bowden J, Davey Smith G. Evaluating the potential role of pleiotropy in Mendelian randomization studies. Hum Mol Genet. 2018;27(R2):R195-r208.

20. Hemani G, Zheng J, Elsworth B, et al. The MR-Base platform supports systematic causal inference across the human phenome. Elife. 2018;7.

21. Bowden J, Hemani G, Davey Smith G. Invited Commentary: Detecting Individual and Global Horizontal Pleiotropy in Mendelian Randomization-A Job for the Humble Heterogeneity Statistic? Am J Epidemiol. 2018;187(12):2681–2685.

22. Moore SC, Lee IM, Weiderpass E, et al. Association of Leisure-Time Physical Activity With Risk of 26 Types of Cancer in 1.44 Million Adults. JAMA Intern Med. 2016;176(6):816–825.

23. Labrecque JA, Swanson SA. Interpretation and Potential Biases of Mendelian Randomization Estimates With Time-Varying Exposures. Am J Epidemiol. 2019;188(1):231–238.

24. Doherty A, Smith-Byrne K, Ferreira T, et al. GWAS identifies 14 loci for device-measured physical activity and sleep duration. Nat Commun. 2018;9(1):5257.

25. Painter JN, O’Mara TA, Marquart L, et al. Genetic Risk Score Mendelian Randomization Shows that Obesity Measured as Body Mass Index, but not Waist:Hip Ratio, Is Causal for Endometrial Cancer. Cancer epidemiology, biomarkers & prevention : a publication of the American Association for Cancer Research, cosponsored by the American Society of Preventive Oncology. 2016;25(11):1503–1510.

26. Swanson SA, Labrecque J, Hernan MA. Causal null hypotheses of sustained treatment strategies: What can be tested with an instrumental variable? Eur J Epidemiol. 2018;33(8):723–728.

27. Vink JM, Boomsma DI, Medland SE, et al. Variance components models for physical activity with age as modifier: a comparative twin study in seven countries. Twin Res Hum Genet. 2011;14(1):25–34.

28. Dowd KP, Szeklicki R, Minetto MA, et al. A systematic literature review of reviews on techniques for physical activity measurement in adults: a DEDIPAC study. Int J Behav Nutr Phys Act. 2018;15(1):15.

29. Folley S, Zhou A, Hypponen E. Information bias in measures of self-reported physical activity. Int J Obes (Lond). 2018;42(12):2062–2063.

